# A Population Level Study on the Determinants of COVID-19 Vaccine Hesitancy at the U.S. County Level

**DOI:** 10.1101/2023.07.12.23292582

**Authors:** Ensheng Dong, Kristen Nixon, Lauren M. Gardner

## Abstract

Multiple COVID-19 vaccines were proven to be safe and effective in curbing severe illness, but despite vaccine availability, uptake rates were relatively low in the United States (U.S.), primarily due to vaccine hesitancy. To better understand factors associated with COVID-19 vaccine hesitancy in the U.S., our study provides a comprehensive, data-driven population-level statistical analysis at the county level. We find that political affiliation, as determined by the proportion of votes received by the Republican candidate in the 2020 presidential election, has the strongest association with COVID-19 vaccine hesitancy. The next strongest association was median household income, which has a negative association. The percentage of Black people and the average number of vehicles per household are also positively associated with vaccine hesitancy. In contrast, COVID-19 infection rate, percentage of Hispanic people, postsecondary education percentage, median age, and prior non-COVID-19 childhood vaccination coverage are other factors negatively associated with vaccine hesitancy. Unlike previous studies, we do not find significant relationships between cable TV news viewership or Twitter misinformation variables with COVID-19 vaccine hesitancy. These results shed light on some factors that may impact vaccination choice in the U.S. and can be used to target specific populations for educational outreach and vaccine campaign strategies in efforts to reduce vaccine hesitancy.

## Introduction

Vaccines are arguably the most effective tool for combating COVID-19, reducing the number of cases, and more critically, severe illness and hospitalization rate, from the disease^1^. One study estimated that the COVID-19 vaccine saved 14.4 million lives globally within one year of its introduction^2^. For the U.S., a study estimated that 240,797 COVID-19 deaths could have been prevented through vaccination from December 12, 2020 to June 30, 2021^3^. As of December 2021, one year since the COVID-19 vaccinations began in the U.S., only 63% of the population completed the primary series of an approved COVID-19 vaccine^4^. In contrast, Canada, Japan, and Italy reached vaccination rates of 70% and above by December 2021^5^. Despite growing evidence for the safety and effectiveness of vaccines, vaccine hesitancy remains influential and is driven by lack of trust in COVID-19 vaccines, concerns about side effects, and lack of trust in government^6^. Better understanding the factors that are associated with vaccine hesitancy is crucial to address this problem.

To investigate potential drivers of vaccine hesitancy, hundreds of existing studies have been conducted using surveys. Aw et al. summarized 97 of these survey-based studies in high-income countries and regions (39 of the articles were specific to the U.S.) and found that factors associated with higher vaccine hesitancy included younger age, females, non-white ethnicity, lower education, lack of recent history of influenza vaccination, lower self-perceived risk of contracting COVID-19, lesser fear of COVID-19, and not having chronic medical conditions^7^. These studies are valuable sources of individual level data and can explore psychological factors that impact hesitancy, but are limited by relatively low sample size and sampling bias. Therefore, population level studies are needed to determine whether these survey findings are generalizable.

Most existing studies at the population level for the U.S. are at the county level and use linear models to examine the relationship between vaccine coverage and demographic features, like income, race, and political affiliation. Multiple studies found political affiliation to be a strong predictor^8,9^. Other demographic features found to be associated with vaccination coverage were socioeconomic status^8,10^, race^8,10^, education level^6,10^, insurance coverage,^8,10^ age^8^, and vehicle access^6,11^. Only two population-level studies incorporated data on information consumption. One study found that more viewership of Fox News during January and February 2020 was associated with lower weekly vaccination uptake between May and June 2021, a relationship that held even when political affiliation was controlled for^12^. Another study found that the percentage of COVID- 19 vaccine-related misinformation shared on Twitter, in addition to increased GOP vote, was negatively associated with vaccine uptake rates^13^.

To analyze COVID-19 vaccine hesitancy and its determinants, our study uses a population-level statistical analysis conducted at the U.S. county level. We define a new variable derived from vaccination uptake rates as a proxy for vaccine hesitancy, specifically, the percent of a county population that did not receive any dose of a COVID-19 vaccine by December 15, 2021^6^, which we refer to as *vaccine hesitant percentage*. This variable is further described and justified in the data section. We implement weighted generalized additive models (GAMs) to identify the relationships between potential determinants and this COVID-19 vaccine hesitant percentage variable, and we include a more comprehensive set of factors potentially influencing vaccine hesitancy than previous work. The choice of variables and the model are described in detail in the following section.

## Data and Methods

This COVID-19 vaccine hesitancy study is based on the latest publicly available data at the county level in the U.S. and uses a weighted generalized additive model (GAM) for the statistical model. The response variable is COVID-19 vaccine hesitant percentage, and the independent variables cover eight categories, including COVID-19 epidemiological data, demographic, socioeconomic, and land use variables, prior non-COVID vaccination behavior, political affiliation, select cable TV channel viewership ratings, and a Twitter misinformation variable (Table 1). The determinants for this study were chosen based on the set of factors previously identified to be associated with vaccine hesitancy in the literature, as well as our own hypotheses about factors that could potentially influence vaccine hesitancy, e.g., COVID-19 burden in a county and prior non-COVID- 19 childhood vaccination rates. Each variable is defined in detail below. The spatial distribution of each variable is visualized in the Supplementary Material (Figure S1), and the correlation matrix is shown in the Supplementary Material (Figure S2).

**Table 1.** Summary statistics for the raw data of the response variable and determinants

| Variable | N | Mean | SD | Source |
| --- | --- | --- | --- | --- |
| **Response variable** |  |  |  |  |
| COVID-19 vaccine hesitant percentage | 3143 | 45.28 | 13.03 | <sup>14</sup> |
| **Demographic data** |  |  |  |  |
| Percentage of Black people | 3143 | 8.72 | 14.11 | <sup>15</sup> |
| Percentage of Hispanic people | 3143 | 9.79 | 13.68 | <sup>15</sup> |
| Postsecondary education percentage | 3143 | 53.68 | 10.72 | <sup>16</sup> |
| Median age | 3141 | 41.43 | 5.42 | <sup>15</sup> |
| **Socioeconomic data** |  |  |  |  |
| Uninsured percentage | 3141 | 9.64 | 5.11 | <sup>15</sup> |
| Average number of vehicles per household | 3141 | 1.98 | 0.24 | 15 |
| Median household income | 3140 | 55707.45 | 13388.63 | 15 |
| **Political affiliation data** |  |  |  |  |
| Republican percentage | 3115 | 64.96 | 16.14 | 17 |
| **Land use data** |  |  |  |  |
| Population | 3143 | 105456.34 | 335760.39 | 15 |
| Population in urban counties | 1133 | 251129.27 | 527897.79 | 15 |
| Population in rural counties | 2010 | 23343.19 | 23994.75 | 15 |
| **COVID-19 epidemiology data** |  |  |  |  |
| Cumulative COVID-19 case rate | 3143 | 16572.79 | 3999.03 | 18 |
| **Non-COVID-19 vaccination behavior** |  |  |  |  |
| MMR vaccination coverage | 3101 | 0.93 | 0.08 | 19 |
| **Information consumption data** |  |  |  |  |
| FNC viewership | 3071 | 0.75 | 1.33 | 20 |
| CNN viewership | 3071 | 0.27 | 0.42 | 20 |
| MSNBC viewership | 3071 | 0.27 | 0.37 | 20 |
| Local news viewership | 3071 | 4.92 | 2.21 | 20 |
| Twitter misinformation percentage | 904 | 0.39 | 0.49 | 21 |

### Data

#### Response Variable

*COVID-19 vaccine hesitant percentage* (VHP) is our chosen response variable. VHP is calculated as the partial vaccination rate (PVR) subtracted from 100%, where the PVR is defined as the percentage of people in a county who had taken at least one dose of Pfizer (Comirnaty) or Moderna (Spikevax)^14,22^. Since the Johnson & Johnson (J&J) vaccine only requires one dose to be fully vaccinated, the PVR excludes individuals who got the J&J vaccine. However, only 3.3% of vaccinations administered were J&J as of December 15, 2021^23^, the effect of excluding J&J vaccinations is relatively negligible. The PVR data used to compute VHP is sourced from Georgetown University’s U.S. COVID-19 Vaccination Tracking website, which primarily relies on CDC data, supplemented with vaccination data from local health departments where CDC data is incomplete^14^. Vaccination data is not available for 69 counties in Alaska, Nebraska, Georgia and Virginia, so these counties were excluded from the analysis.

Critically, the proposed definition of VHP relies on vaccination uptake rates (more specifically, the complement to the rate) as a proxy for vaccine hesitancy behavior. While this is not a perfect substitute, the choice is further justified based on a previous survey-based study that showed vaccine uptake is strongly correlated with vaccine hesitancy^6^. Further, vaccine uptake rates reflect real world vaccination behavior at the population level, in contrast to vaccine hesitancy surveys which are available for a subset of locations around the U.S. and suffer from sampling biases. Additional implications of this decision are further discussed in the limitations section of this work.

#### Explanatory Variables

##### Demographic and socioeconomic data

All demographic and socioeconomic variables are sourced from publicly available datasets at the county level, from the U.S. Census Bureau’s 2020 Decennial Census^15^ and U.S. Department of Agriculture (USDA) Economic Research Service^16^. The percentage of Black people and the percentage of Hispanic people represent the self-identified proportion of those races in each county. Postsecondary education percentage is measured as the percentage of adults with educational attainment more advanced than completing high school. The uninsured percentage is the percentage of people who reported not having health insurance. Additional metrics include median age, average number of vehicles per household and the median household income.

##### Political affiliation data

The political affiliation variable, defined as the percentage of voters who chose Donald Trump as their presidential candidate during the 2020 presidential election, is sourced from the MIT Election Data and Science Lab^17^. It is hereby referred to as “Republican percentage (%).”

##### COVID-19 epidemiology

In efforts to explore whether a county that experienced more burden from COVID-19 may be more willing to adopt preventative measures such as vaccination, we incorporate a variable to capture a county’s historical COVID-19 infection rate. Specifically, to measure historical COVID-19 burden, we use the cumulative number of COVID-19 cases per 100,00 people as of December 15, 2021 from the Johns Hopkins University Center for Systems Science and Engineering (CSSE) COVID-19 GitHub^18^. In order to remove outliers, values that were more than 4 standard deviations above the mean were excluded.

##### Non-COVID-19 vaccination behavior

As MMR vaccination coverage is an indicator of vaccine acceptance before COVID-19, we hypothesize that higher (pre-pandemic) MMR vaccine uptake rates may be associated with higher COVID-19 vaccine coverage. To test this hypothesis, we incorporate a variable in this analysis that is based on the MMR vaccination coverage rates of children in kindergarten in 2019, which we published in a previous study^19^.

##### Information consumption

A set of variables intended to capture the potential role of information consumption on vaccine choice includes four television viewership rating variables and a Twitter misinformation variable. The county level viewership ratings (RTG) % for four major channels, namely FNC (Fox News Channel), CNN (Cable News Network), MSNBC (Microsoft National Broadcasting Company), and local news, are sourced from Nielsen Media, where RTG is measured by the estimated percentage of households tuned to a specific viewing source, e.g., news channel. The four viewership variables were computed as the average of the monthly viewership ratings for each channel from February to November 2021. January 2021 data were excluded due to anomalies caused by the January 6^th^ U.S. Capital Attack. Nielsen data is not available for several counties in Virginia and Alaska and all counties in Hawai’i, so these 72 counties are excluded from the analysis. The analysis also excludes outliers, defined as those counties with cable viewership values that are more than 4 standard deviations away from the mean. Additionally, within the model each of the cable TV viewership variables was standardized to a mean of 0 and standard deviation of 1, to provide a more interpretable understanding of the relative position of each county’s rating.

Another information consumption variable included in the model is the Twitter misinformation variable. This variable is intended capture the prevalence of COVID-19 vaccine misinformation in circulation on Twitter during a time that likely influenced behavior during the study period. The variable is based on a previous study by Pierri et al., who provided a variable that is representative of the percent of COVID-19 vaccine-related tweets that contain links to low credibility sources at the county level^13,21^. This variable has some limitations, as it is based on only the set of twitter accounts that can be geolocated. To ensure a large enough sample size for a reliable estimate, counties with less than 50 geolocated accounts are not included, which results in a data set that includes 855 counties. An analogous data set is also available with a minimum of 10 and 100 geolocated accounts, but we opted to use the cutoff of 50 to balance having a more representative sample size of accounts per county with the number of counties we can include in our analysis. Due to the limited number of counties that this data is available for, a separate sub-analysis is conducted that includes this variable (Figure 2).

##### Land use data

Various land-use variables are sourced from the U.S. Census Bureau, namely the population size and the number of residents in rural or urban areas for each county^24^. These variables are used to cluster counties for the sub-analyses, further described in the methods section. For the cluster-based analysis we categorize counties into mutually exclusive sets based on 1) population quartiles and 2) a binary rural or urban classification. For the binary classification, a county is classified as rural if the majority of the population is designated to live in areas classified as rural and otherwise classified as urban.

### Statistical Models

We use a Generalized Additive Model (GAM) to explore the relationship between each county’s vaccine hesitant percentage and the aforementioned variables. GAMs provide a balance between model complexity and interpretability, and critically, they can reflect the relative importance of different features^25,26^. Specifically, GAMs model the response variable as the sum of unknown smooth functions of covariates, and unlike Generalized Linear Models (GLMs), GAMs offer flexibility in capturing nonlinearity within variables while accounting for associations among them. Thus, the nuanced nature of the relationships between these variables and the response variable, vaccine hesitant percentage, are better captured by GAMs than simpler linear models.

#### Primary Model

The proposed GAM is fitted to the vaccine hesitant percentage as the response variable, which is assumed to have a Gaussian distribution, and a log link. REML (restricted maximum likelihood) is used to estimate smoothing parameters, which returns relatively reliable and stable results. Specifically, the model in our primary analysis has the following form:

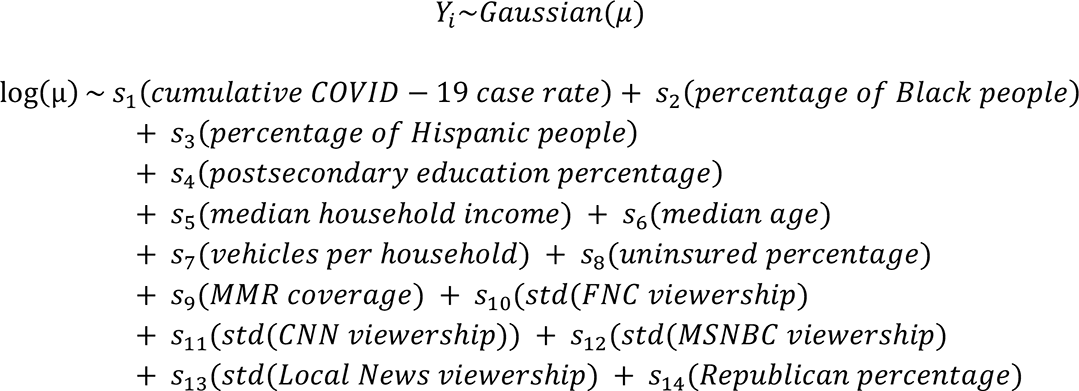

where *Y_i_* denotes the vaccine hesitant percentage for each county *i*. The model is a sum of smooth functions *s_i_*, and each smooth function consists of a number of basis functions (*k*). Sensitivity analysis that varies the number of basis functions was conducted. A value of *k*=3 for each smooth function was found to provide the optimal balance between preventing both underfitting and overfitting of the model and maximizing interpretability of the results. Additionally, the GAM model is weighted to prevent the highly imbalanced county population distribution from skewing the results. The weight is computed by normalizing each county’s population by the average county population, taking a log transformation to adjust for the skewness. The weight implemented in the primary analysis is defined as:

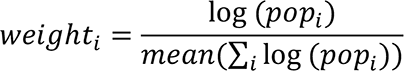

where *i* is the county index. The primary model is run for 2,885 counties (reduced from the full set of counties due to missing data and data quality issues referenced previously).

As noted previously, the Twitter misinformation variable, *s*_15_(*Twitter misinformation*), is only available for 885 counties, and is therefore run as a separate model using the same general function and weights as the primary model, but with the additional determinant included.

#### Sensitivity Analysis

In addition to the primary model presented above, we conduct sub-analyses to further investigate the relationship between vaccine hesitant percentage and its associated factors, based on modeling subsets of counties independently. Two cluster-based sub-analyses are motivated by the hypothesis that there exists heterogeneity among U.S. residents with respect to various factors not accounted for in this study, such as perceived COVID-19 disease risks and burden, trust in scientific and government institutions, public health policy and policy compliance, which might lead to differences in the associations of the modeled factors and the vaccine hesitant percentage variable of interest. To test this hypothesis, separate models are fit for subsets of the counties based on underlying land use patterns or population levels. Specifically, the sub-analyses are as follows:

1. Land-use cluster-based analysis: Counties are clustered into two groups based on their primary land use pattern, namely as urban or rural counties. Two independent weighted GAM models are run, one for each group. The rural model includes 1,835 counties, and the urban model includes 1,050 counties.
2. Population cluster-based analysis: Counties are grouped into quantiles based on their population size. Four independent GAM models are generated, one for each distinct quantile. The respective models contain 664, 721, 739, and 761 counties ranging from the smallest to largest population size groups. GAMs are implemented without weights for each group in this sub-analysis, because the weighting is based on population size.

We evaluate the goodness-of-fit by conducting a diagnostic analysis for each model and sub-model. These evaluations include the Q-Q plots, histograms of residuals, mapping of residual values versus predicted values, and mapping of response against fitted values. The diagnostic analysis outcomes for the primary model are presented in the Supplementary Material (Figure S4). The concurvity in the primary model is also measured to ensure pairwise values remain below 0.8 and avoid cases in which one variable is a smooth function of another. The outcomes of the diagnostic analysis demonstrated consistency in fit and performance across all models.

## Results

The GAM results are presented for each analysis: 1) the primary model for 2,885 counties in the U.S. (Figure 1), 2) the sub-model that includes a Twitter misinformation variable for 855 counties (Figure 2), 3) the land-use cluster-based analysis with separate models for counties classified as urban or rural (Figure 3), and 4) the population cluster-based analysis with separate models for counties grouped based on population size in the Supplementary Material (Figure S5).

**Figure 1.**
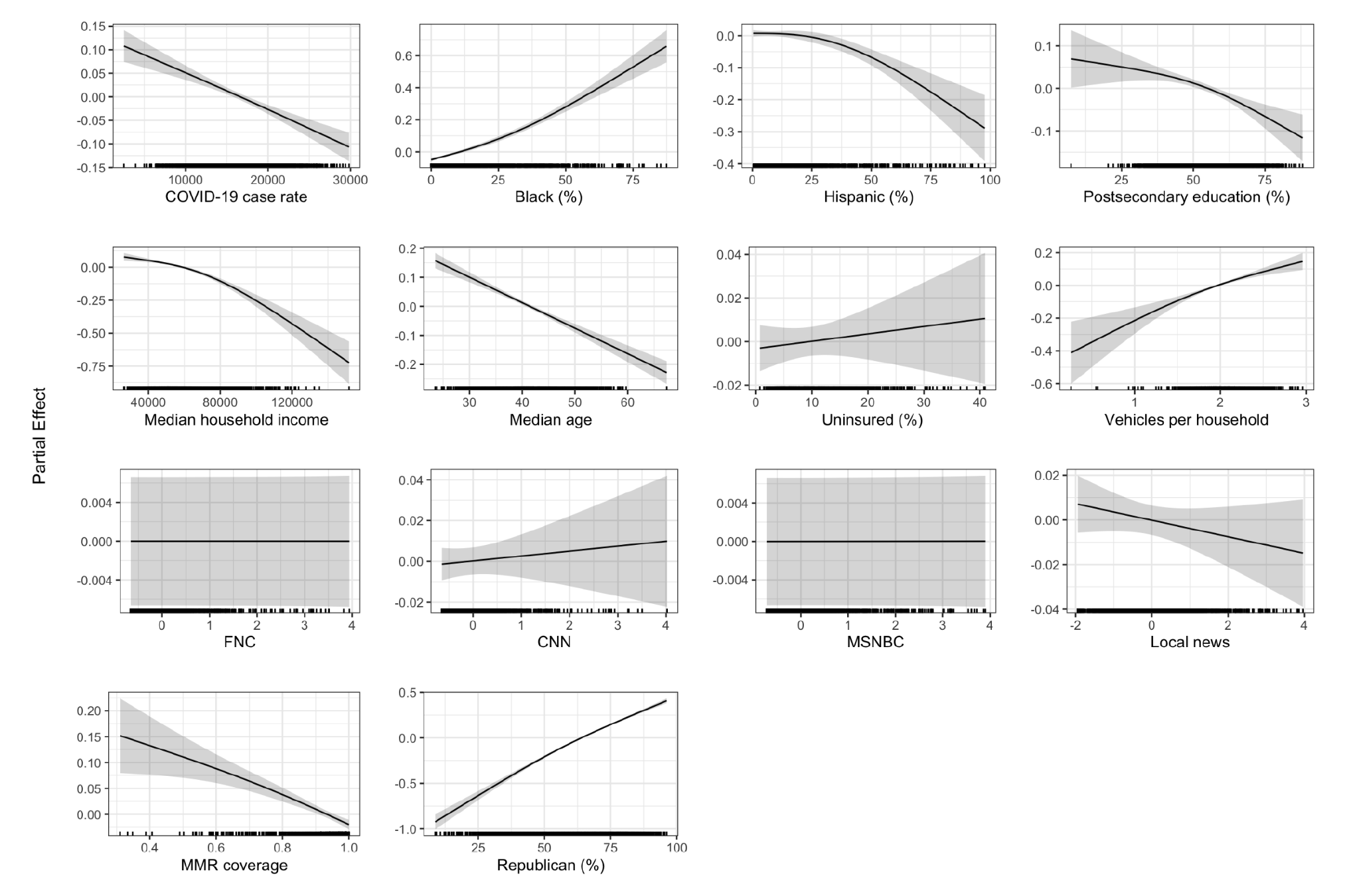
GAM results for the primary model. The shaded regions in each curve refer to the 95% confidence intervals, and the rugs at the bottom of each subplot indicate the distribution of each determinant.

**Figure 2.**
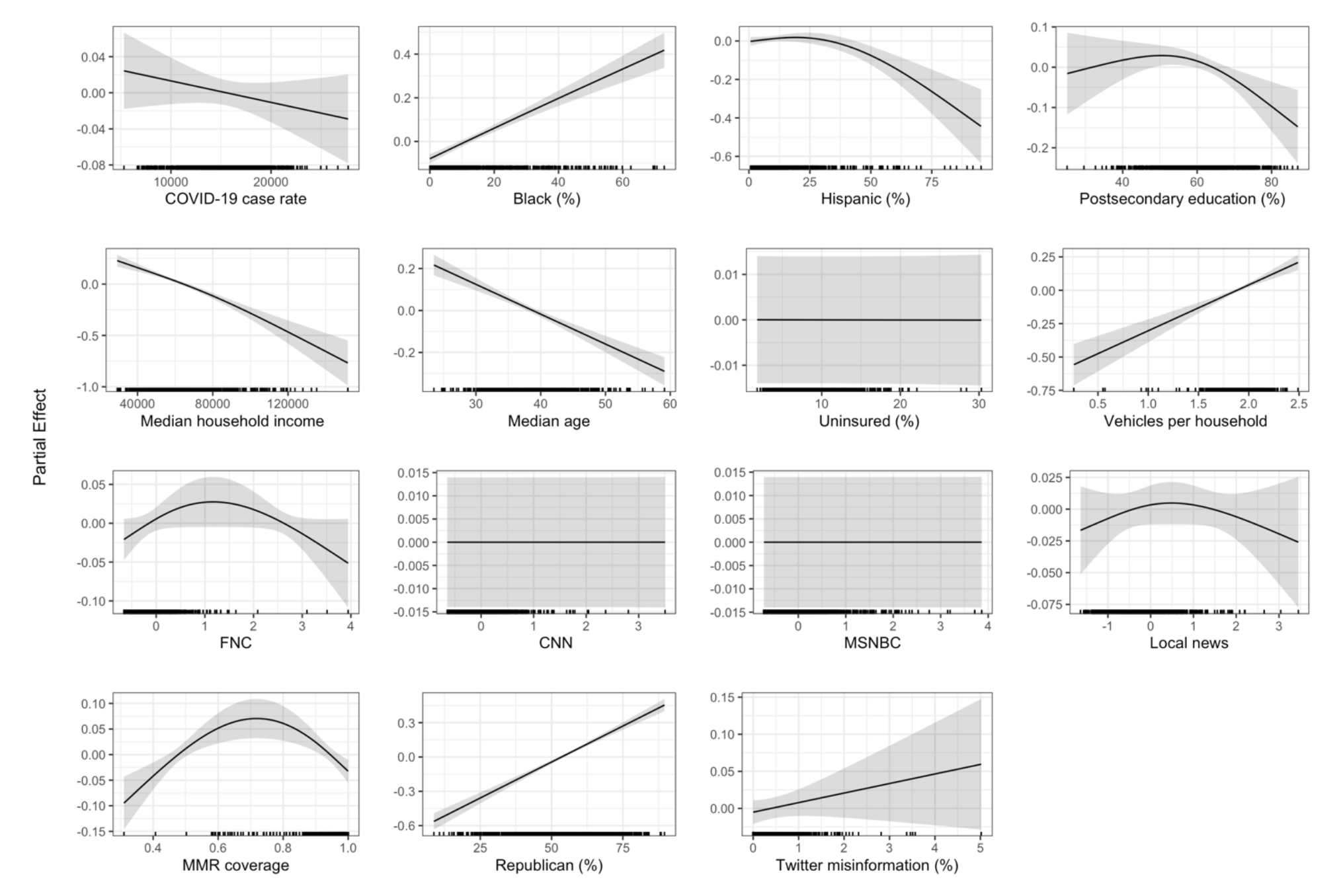
Results of the generalized additive models with Twitter misinformation (%) with 855 counties in the model. The shaded regions in each curve refer to the 95% confidence intervals, and the rug at the bottom of each subplot indicates the distribution of each determinant.

**Figure 3.**
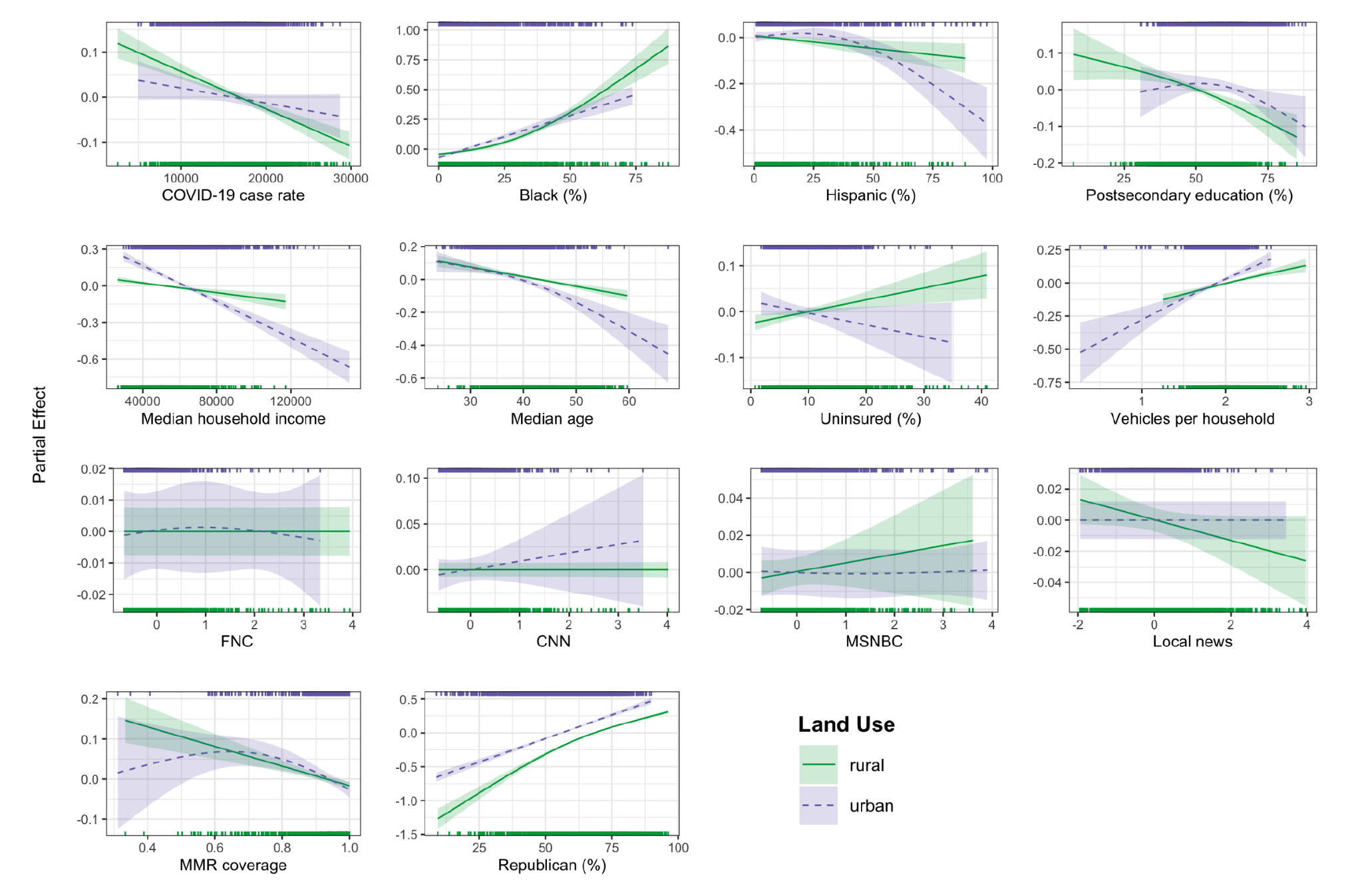
Results of the generalized additive models clustered by land use (i.e., rural counties vs. urban counties). The shaded regions in each curve refer to the 95% confidence intervals, and the rugs at the top and bottom of each subplot indicate the distribution of each determinant by cluster.

### Primary model of vaccine hesitancy and associated factors in the U.S

Figure 1 contains the GAM results for the primary model that includes the majority of U.S. counties. The factor with the strongest positive association with COVID-19 vaccine hesitant percentage in our model is Republican (%), i.e., the percentage of voters choosing Donald Trump as their presidential candidate during the 2020 presidential election. Two other determinants positively associated with vaccine hesitant percentage are the percentage of Black people and the average number of vehicles per household, however the associations are not as strong as the Republican (%). Multiple variables are found to be negatively associated with vaccine hesitant percentage, namely cumulative COVID-19 case rate, percentage of Hispanic people, postsecondary education percentage, median household income, median age, and MMR vaccination coverage, although the strength of the associations varies, with the strongest negative association appearing for median household income. In contrast, the uninsured percentage and the cable TV viewership variables do not show a statistically significant relationship (*p* < 0.05) with vaccine hesitant percentage in the primary model (see Table S1 in the Supplementary Material). Results with a unified y-axis range are shown in the Supplementary Material (Figure S3), which further illustrates the strong relative association of Republican percentage compared to other determinants.

### Sub-model incorporating Twitter misinformation rates

Figure 2 presents results from the sub-model for 855 counties that includes the Twitter misinformation variable from Pierri et al^13^, which represents the proportion of COVID-19 vaccine-related tweets sharing low credibility sources in a county. Based on the model results, and in contrast to previous work^13^, the Twitter variable does not have a significant association with vaccine hesitant percentage. The results from both the primary model and this sub-analysis are broadly consistent, with the only differences being that for the model with twitter misinformation, the cumulative COVID-19 case rate is not significant, Fox News viewership has a significant inverted U-shaped trend, and the postsecondary education percentage has a slight inverted U-shaped trend. The trends for COVID-19 case rate and postsecondary education are consistent with our findings in urban counties and counties with larger populations (see cluster-based analysis), which reflects that the sample of counties that have Twitter misinformation data are biased towards larger urban counties. The inverted shape of the Fox News trend is driven by a very small number of counties with viewership values greater than 2, while the positive slope is consistent with the vast majority of counties included in the model.

### Land-use cluster-based sensitivity analysis

Figure 3 shows the results of the land use cluster-based sensitivity analysis, which fits separate models for rural and urban counties in efforts to determine if the different experiences of urban versus rural residents’ results in different factors and associations with the vaccine hesitant percentage. The sub-models reveal the same significant variables as the primary model with few exceptions. For rural counties there is a positive association between the uninsured percentage and vaccine hesitancy, compared to no significant relationship in the primary model and for urban counties. The rural counties also have a negative association between local news viewership and vaccine hesitancy, while the primary and urban counties have no significant relationship between these variables. When comparing the urban and rural counties, rural counties reveal a stronger negative association between cumulative COVID-19 case rate and vaccine hesitancy, while urban counties show stronger trends for the negative association between percentage of Hispanic people and vaccine hesitancy, and the negative association between median household income and vaccine hesitancy. The trends for postsecondary education and MMR vaccination coverage are more complex in urban counties, forming a more uncertain, slightly inverted U-shaped trend, compared to a more linear trend for rural counties. The slightly positive trend for counties with MMR coverage less than 60% is driven by only three counties, and as evidenced by the wide uncertainty interval, is not a significant trend.

### Population cluster-based sensitivity analysis

Results for the population cluster-based sensitivity analysis is presented in the Supplementary Material (Figure S5). Four independent models are fit for the sets of counties clustered by population quartile to further assess the robustness of the associations identified in the primary and sub-analysis. The results from the population sub-models are consistent with the models for urban and rural counties. Specifically, the negative association between cumulative COVID-19 case rate and vaccine hesitant percentage is higher in counties with smaller population, the percentage of Hispanic people’s negative association is stronger in counties with larger population, local news viewership only has a significant negative association in smaller counties, and postsecondary education percentage has a stronger negative association in smaller counties. For MMR coverage, the smallest quartile shows no relationship with vaccine hesitancy, the middle two quartiles have a clear negative association, and the largest quartile has an inverted U-shaped trend. Like the urban counties model, the positive association in the largest quartile when MMR coverage is less than 60% is driven by a very small number of counties with large population size and low MMR vaccination coverage.

## Discussion

Across all models presented, political affiliation, namely the percentage of voters who voted for Donald Trump in the 2020 presidential election, has the strongest association with COVID-19 vaccine hesitant percentage in the U.S. This result is consistent with previous studies at the individual level^27,28^ and at the population level^8,9,13^.

For demographic variables, a high percentage of Black people is positively associated with vaccine hesitant percentage, while a high percentage of Hispanic people is negatively associated with vaccine hesitant percentage. These results are consistent with previous studies^8,10,13^. Frisco et al. found evidence that the reason for more vaccine hesitancy among Black people is due to the legacy of racism Black Americans have faced in medicine and medical research, while the same study found evidence that decreased vaccine hesitancy among Hispanic people is due to Hispanic people experiencing higher burden from COVID-19^29^.

Other demographic and socioeconomic factors negatively associated with vaccine hesitant percentage include median age, median household income, and postsecondary education percentage, while the average number of vehicles per household has a positive association. These results are also consistent with previous work. Older people were able to access the vaccine earlier and are more susceptible to COVID-19, which likely decreased their vaccine hesitancy^8^. Higher levels of educational attainment^6,10^ and higher income^8,10^ were associated with decreased vaccine hesitancy. While previous work found that greater insurance coverage is broadly associated with higher vaccine uptake^8,10^, our results identified this relationship only in rural counties. This may reflect that health insurance was more influential for certain subpopulations with more limited access to vaccines and healthcare in general, as is the case in rural counties^30^. More vehicles per household was associated with increased vaccine hesitancy, which is consistent with previous work that shows higher vaccine coverage to be associated with lower percentages of households with a vehicle^6,11^. This finding seems counterintuitive since vehicle access suggests easier access to vaccines. However, urban counties have less vehicles on average than rural counties, but urban residents typically enjoy closer proximity to vaccine centers and transit options other than private vehicles. Therefore, this relationship may be capturing confounding factors such as urban residents typically having easier access to vaccines and less vaccine hesitancy.

Our study found that higher COVID-19 case rates are associated with lower vaccine hesitancy. We hypothesize that in counties that experienced a higher historical case burden of COVID-19, individuals were more aware of the risks of COVID-19 and thus more willing to seek out preventative measures like vaccination. However, this relationship was weaker in urban counties and was insignificant in our model with a Twitter misinformation variable, which is biased towards counties with larger populations. In most cases, historical burden of COVID-19 appears to influence the perceived risk versus the reward of vaccination and encourage uptake, except in counties with larger populations, in which other determinants were more important.

Results from our analysis revealed a negative association between prior non-COVID-19 vaccination behavior, measured by MMR vaccination coverage, and COVID-19 vaccine hesitancy. This relationship was stronger in rural counties than urban counties. The MMR vaccination coverage is an indicator of vaccine acceptance before COVID-19, which is based on vaccination coverage of children in kindergarten, and reflects their parents’ acceptance of recommended childhood vaccinations. That higher MMR vaccination coverage was associated with lower COVID-19 vaccine hesitant percentage suggests that vaccine hesitancy in the U.S. was not necessarily specific to COVID-19 vaccines, and that the same populations that were historically hesitant towards recommended childhood vaccinations (for their children) were also hesitant towards COVID-19 vaccination (for themselves). These results suggest that successful vaccination campaigns can help increase vaccination uptake more broadly, but they must address the more complex issue of general hesitancy towards vaccines, rather than just concerns around a specific disease.

Previous work found that cable news viewership had a significant relationship with vaccine hesitancy. However, results from our more comprehensive model, which includes a broader scope of variables that impact vaccine hesitancy, did not indicate consistent and significant relationships for these cable news viewership variables. The only significant relationships with vaccine hesitant percentage found were for local news viewership (negative association) in rural counties and Fox News viewership (uncertain, inverted U-shaped trend) in the sub-model with a Twitter misinformation variable. In contrast, a previous study on viewership of cable TV found evidence of causality between Fox News viewership and lower weekly vaccine uptake between May and June 2021, a relationship that holds when controlling for self-reported political affiliation^11^. In addition to considering a more limited range of variables, the study focuses on a smaller timespan and defines political affiliation using the Gallup Polling Series 2012-2019, rather than 2020 presidential election voting.

Our study also found no significant relationship between the Twitter misinformation variable provided by Pierri et al. and COVID-19 vaccine hesitancy, which also contrast previous findings^13^. Pierri et al. found that this Twitter misinformation variable was the most significant predictor for vaccine coverage at the state level, followed by political affiliation^13^, however, at the county level political affiliation was shown to be a stronger predictor of vaccine coverage than Twitter misinformation. Our results reveal that in a more comprehensive framework at the county level, this Twitter misinformation variable does not have a significant effect on the vaccine hesitant percentage relative to the other factors modeled. However, our study does not necessarily indicate that information consumption does not impact vaccine hesitancy, but more likely that we do not yet have data that accurately captures information consumption patterns that influence health-related behaviors.

There are several limitations of this study. First, we calculated COVID-19 vaccine hesitant percentage as the partial vaccination rate subtracted from 100%, which reflects the absence of vaccine uptake, not necessarily people’s attitudes towards vaccines. Vaccination uptake is affected by mandatory requirements from local health departments, schools, or workplaces, which may override a person’s preference with respect to COVID-19 vaccination. In addition, the use of vaccination uptake as a proxy for vaccine hesitancy is not perfect and prohibits our ability to separate out non-hesitancy factors such as eligibility and accessibility, which may also impact vaccination uptake. However, to minimize the effect of these other factors, we calculate our vaccine hesitancy variable based on vaccination uptake as of December 15, 2021. This time cutoff is eight months after the vaccine became available to all adults in the U.S., at which time the impact of the non-hesitancy related factors such as access would be relatively negligible. Second, attitudes toward vaccines evolved over time due to both scientific and anecdotal influences. Our study considers data up until December 2021 and does not account for variations over time. Third, our findings are only applicable to the U.S. Fourth, our analysis is conducted at the population level, therefore our results do not reflect any individual level findings. Fifth, GAMs are vulnerable to overfitting, especially to outliers, as shown in the MMR plot for urban counties and the Fox News trend in the model with Twitter misinformation. To counteract this tendency, we have removed outliers for variables when necessary and explicitly discussed when trends are not significant due to these issues. Finally, our analysis is driven by statistical correlations and therefore cannot make any claims about casual relationships.

## Conclusion

This study examines about 3,000 U.S. counties covering over 300 million people to analyze the determinants associated with COVID-19 vaccine hesitancy in the U.S. In spite of the inclusion of multiple variables that would intuitively influence vaccination-related decision-making, such as historical COVID-19 burden, non-COVID-19 vaccination uptake patterns, and a variable on the prevalence of COVID-19 vaccine misinformation on Twitter, we found political affiliation, defined by voting rates for Donald Trump in the 2020 election, to be the most strongly associated variable with vaccine hesitancy. These findings highlight the harms of the politicization of COVID-19 in the U.S. and its influence on public health decision-making, most critically, during the time period when vaccination was the most powerful tool for fighting COVID-19, and the burden from COVID-19 was at a peak. In addition, our results identified the second strongest determinant of vaccine hesitancy, behind political affiliation, to be median income, which underscores the longstanding role of inequality in the U.S. driving disparate health outcomes. We also found significant associations with race, education level, and vehicle access. That these demographic and socioeconomic factors are associated with vaccine hesitancy, along with many other public health outcomes of interest, illustrates the ongoing role of these factors in driving the inequitable burden of disease. In summary, this study builds on previous work with the goal of understanding what factors are associated with vaccine hesitancy in the U.S., in order to guide future efforts to increase vaccination uptake for COVID-19, and other vaccine preventable diseases.

## Data Availability

The data utilized in the study, along with the corresponding code, have been published on GitHub: https://github.com/CSSEGISandData/covid19_vaccine_hesitancy_study. However, in compliance with the redistribution rules of Nielsen, the cable TV viewership data was excluded from the shared dataset.

## Author contributions

Concept and design: L.G., E.D., Data collection: E.D., Code and figures preparation: E.D., Data analysis: E.D., K.N., L.G., Methodology: L.G., E.D., Writing: E.D., K.N., L.G. Supervision and funding acquisition: L.G. All authors reviewed the results and approved the final version of the manuscript.

## Competing interests statement

The authors declare no potential conflicts of interest.

## Supplementary material for

**Figure S1.**
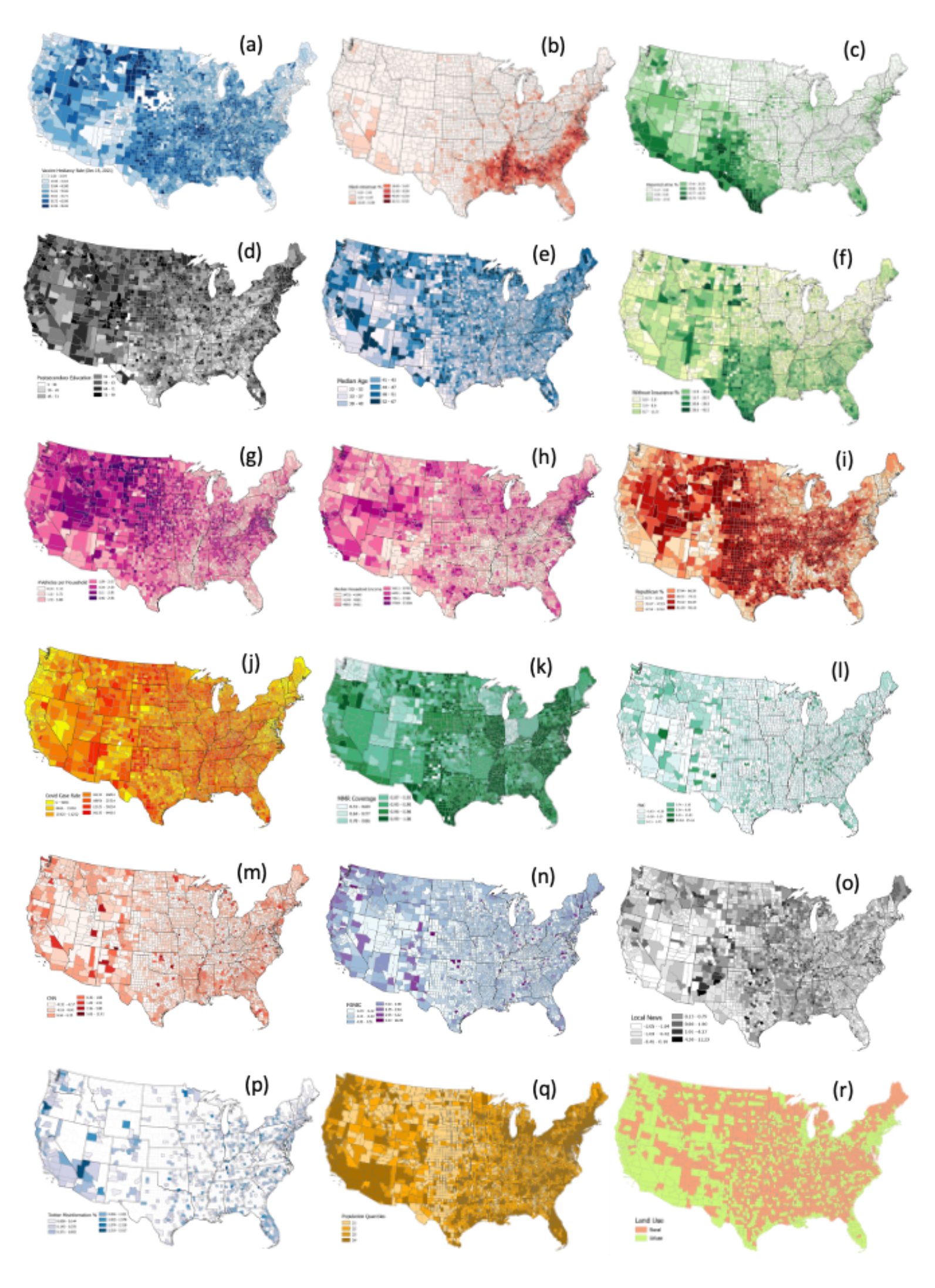
Spatial distributions of all data used in this study. (a) COVID-19 vaccine hesitant percentage; (b) Percentage of Black people; (c) Percentage of Hispanic people; (d) Postsecondary education percentage; (e) Median age; (f) Uninsured percentage; (g) Average number of vehicles per household; (h) Median household income; (i) Republican percentage; (j) COVID-19 case rate; (k) MMR vaccination coverage; (l) FNC viewership; (m) CNN viewership; (n) MSNBC viewership; (o) Local news viewership; (p) Twitter misinformation percentage; (q) Population quantiles; (r) Land use type.

**Figure S2.**
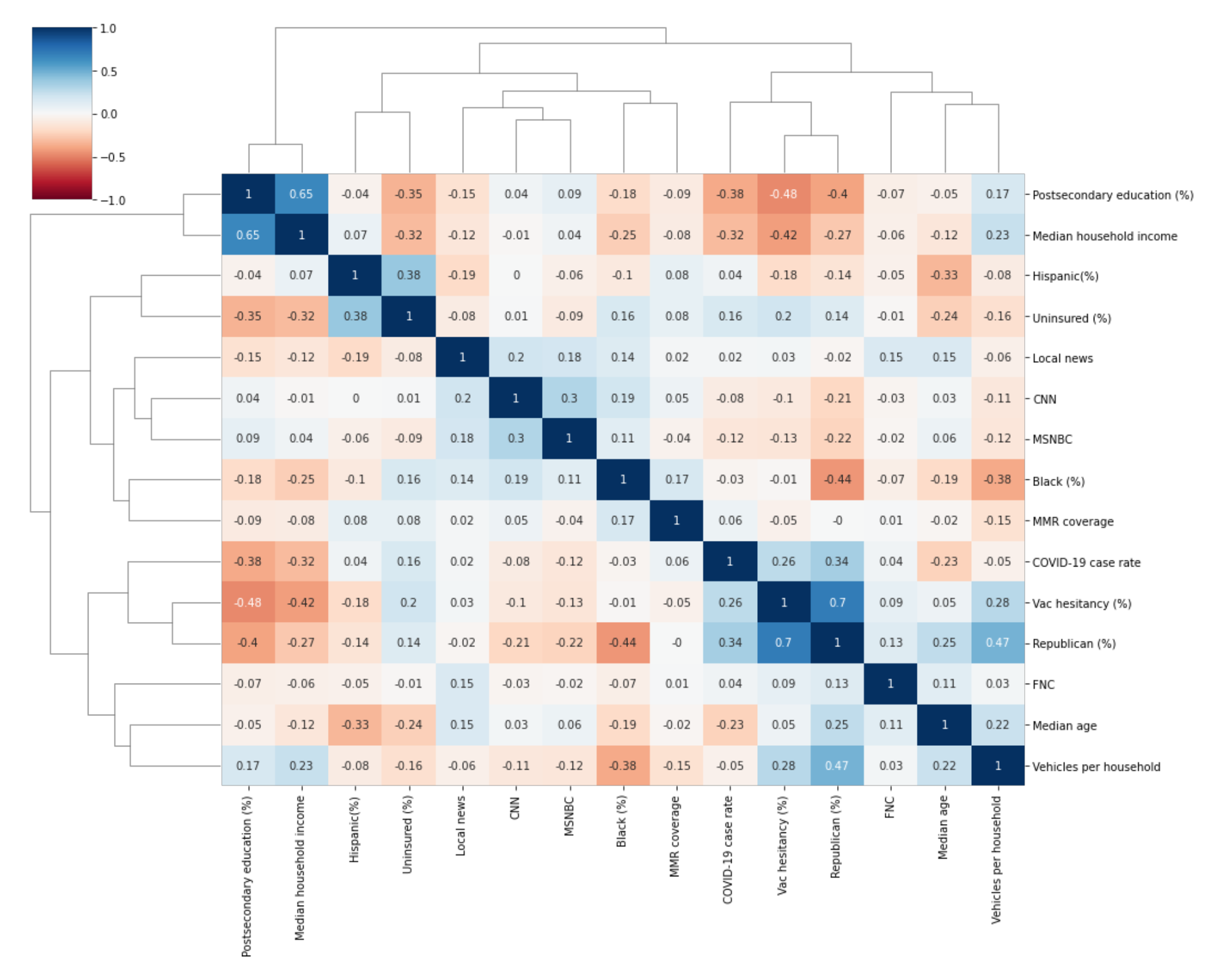
Correlation matrix among the outcome and the determinants for data in the primary model.

**Figure S3.**
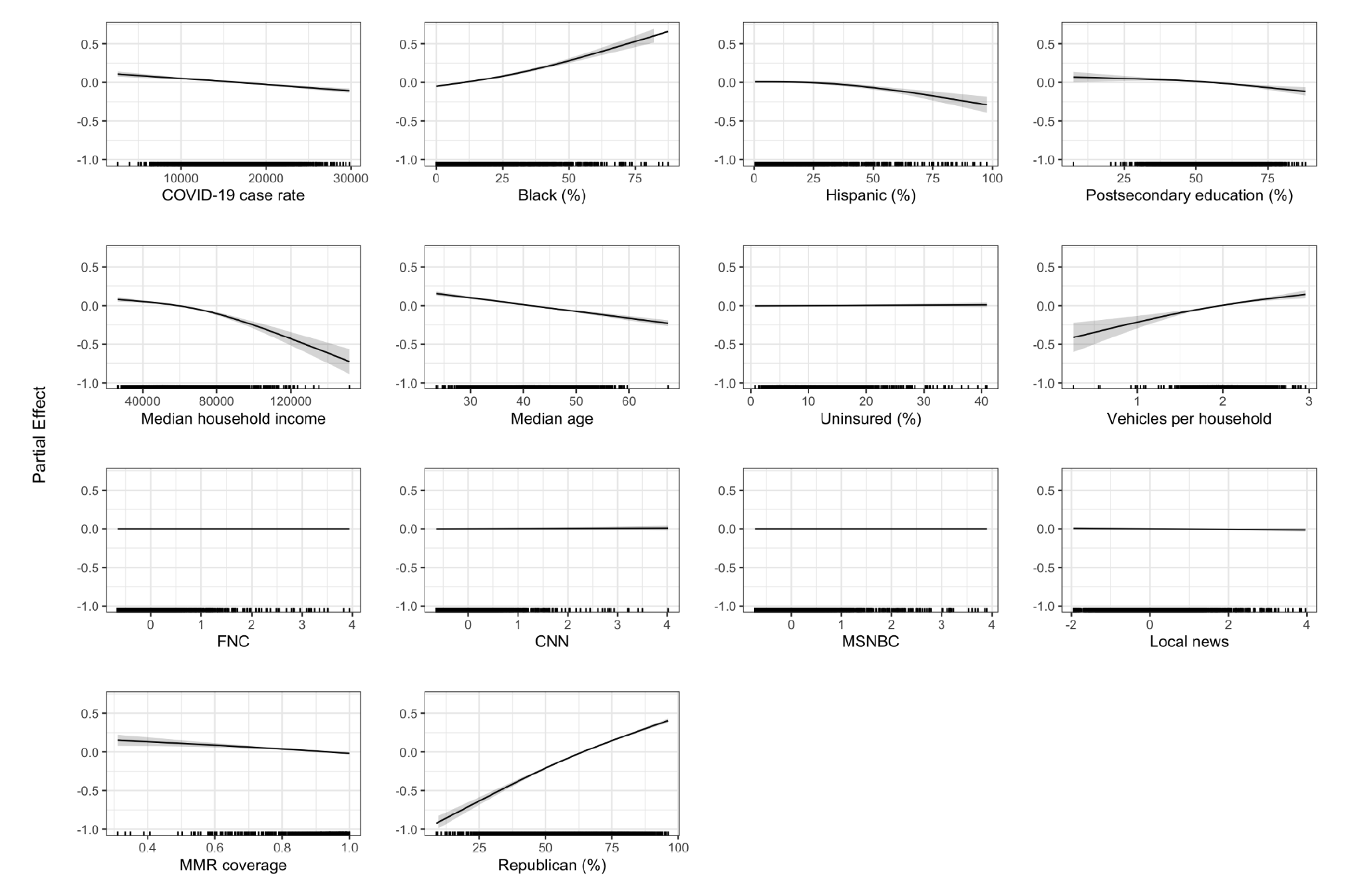
GAM results for the primary model with unified y-axis range. The shaded regions in each curve refer to the 95% confidence intervals, and the rug at the bottom of each subplot indicates the distribution of each determinant.

**Figure S4.**
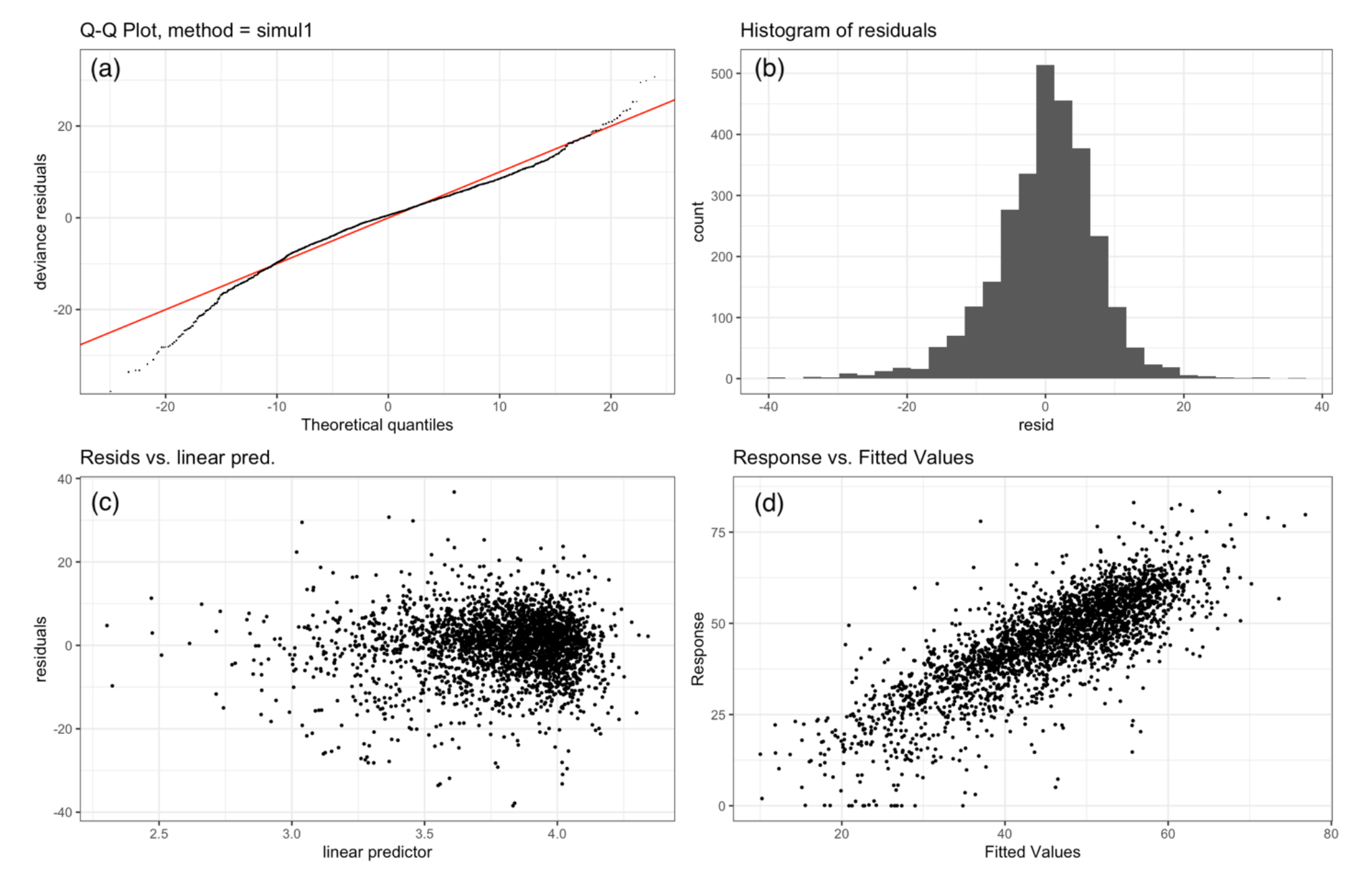
Model evaluations for the GAM result of the primary model. (a) Q-Q plot; (b) histogram of residuals; (c) plot of residual values versus predicted values; (d) plot of response against fitted values.

**Figure S5.**
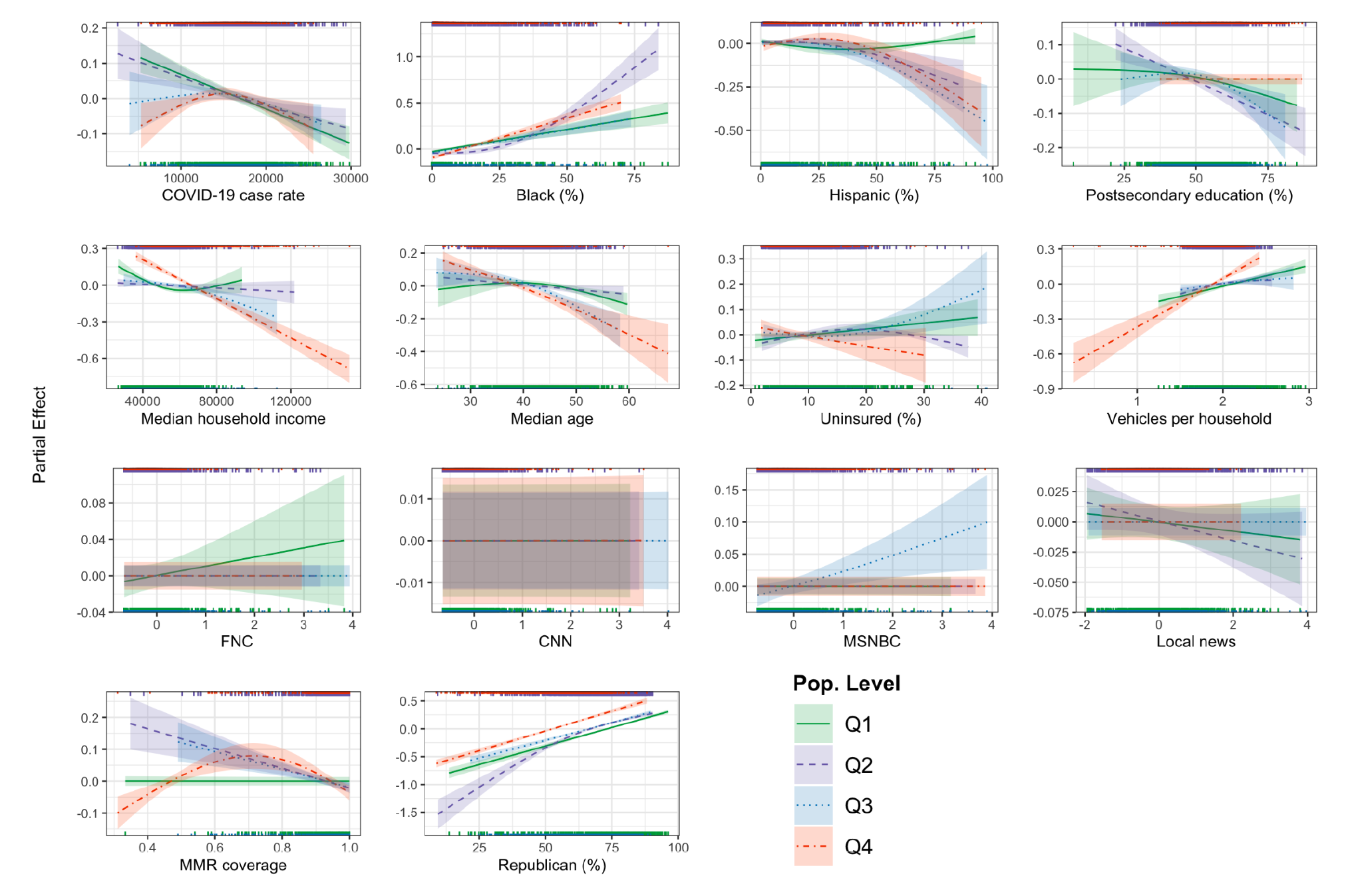
Results for the population cluster-based sensitivity analysis. Counties are clustered into quartiles based on population size, with thresholds at 10,830 (similar size to Sussex, VA), 26,000 (similar size to Staunton, VA), and 68,000 (similar size to Madison, NY). Q1 is the smallest quartile, and Q4 the largest. The shaded regions in each curve refer to the 95% confidence intervals, and the rugs at the top and bottom of each subplot indicate the distribution of each determinant.

**Table S1.** P-values for significance of smooth terms in each GAM. Values less than 0.05 are marked with an asterisk (*).

| Variables | Primary Model | Rural Counties | Urban Counties | Population Q1 | Population Q2 | Population Q3 | Population Q4 | Twitter Misinfo |
| --- | --- | --- | --- | --- | --- | --- | --- | --- |
| COVID-19 case rate | < 2e-16 * | < 2e-16 * | 0.0422 * | < 2e-16 * | 1.34e-05 * | 0.00547 * | 0.008819 * | 0.11614 |
| Percentage of Black people | < 2e-16 * | < 2e-16 * | < 2e-16 * | < 2e-16 * | < 2e-16 * | < 2e-16 * | < 2e-16 * | < 2e-16 * |
| Percentage of Hispanic people | < 2e-16 * | 0.00264 * | 4.14e-06 * | 0.033242 * | 0.001342 * | 3.63e-05 * | 0.000141 * | 1.5e-05 * |
| Postsecondary education | < 2e-16 * | < 2e-16 * | 0.02 * | 0.024678 * | 3.32e-06 * | 0.00404 * | 0.423629 | 0.00178 * |
| Median household income | < 2e-16 * | 2.17e-05 * | < 2e-16 * | 1.64e-06 * | 0.110638 | 3.73e-05 * | < 2e-16 * | < 2e-16 * |
| Median age | < 2e-16 * | < 2e-16 * | < 2e-16 * | 0.000939 * | 0.017459 * | < 2e-16 * | < 2e-16 * | < 2e-16 * |
| Uninsured percentage | 0.217 | 0.00114 * | 0.066 | 0.026748 * | 0.014393 * | 0.01699 * | 0.045869 * | 0.29917 |
| Vehicles per household | < 2e-16 * | < 2e-16 * | < 2e-16 * | 5.48e-07 * | 0.000404 * | 0.01160 * | < 2e-16 * | < 2e-16 * |
| Fox News viewership | 0.975 | 0.7621 | 0.2869 | 0.133697 | 0.389378 | 0.58551 | 0.804054 | 0.03507 * |
| CNN viewership | 0.236 | 0.48825 | 0.1758 | 0.578328 | 0.816158 | 0.54402 | 0.36291 | 0.77838 |
| MSNBC viewership | 0.454 | 0.15819 | 0.3012 | 0.377947 | 0.801062 | 0.00291 * | 0.769641 | 0.94142 |
| Local news viewership | 0.105 | 0.03535 * | 0.9412 | 0.187104 | 0.058216 | 0.56133 | 0.845971 | 0.14927 |
| MMR coverage | < 2e-16 * | 9.18e-07 * | 3.97e-05 * | 0.359801 | 7.92e-06 * | 4.12e-05 * | 3.05e-05 * | 8.8e-05 * |
| Republican percentage | < 2e-16 * | < 2e-16 * | < 2e-16 * | < 2e-16 * | < 2e-16 * | < 2e-16 * | < 2e-16 * | < 2e-16 * |
| Twitter misinformation % | n/a | n/a | n/a | n/a | n/a | n/a | n/a | 0.09348 |

## Notes

### Competing Interest Statement

The authors have declared no competing interest.

### Funding Statement

This study did not receive any funding.

